# The spread and burden of the COVID-19 pandemic in sub-Saharan Africa: comparison between predictions and actual data and lessons learned

**DOI:** 10.1101/2022.05.04.22274692

**Authors:** Christophe Dongmo Fokoua-Maxime, Yahia Bellouche, Dillonne Ngonpong Tchigui-Ariolle, Tchana Loic Tchato-Yann, Simeon Pierre Choukem

## Abstract

**Introduction:** Sub-Saharan Africa (SSA) was predicted to be severely affected by the coronavirus disease 2019 (COVID-19) pandemic, but the actual data seem to have contradicted these forecasts. This study attempted to verify this observation by comparing predictions against actual data on the spread and burden of the COVID-19 pandemic in SSA.

**Methods:** Focused on the period from March 1^st^ to September 30^th^, 2020, we compared (1) the predicted interval dates when each SSA country would report 1 000 and 10 000 COVID-19 cases, to the actual dates when these numbers were attained, as well as (2) the daily number of predicted versus actual COVID-19 cases.

Further, we calculated the case fatality ratio of the COVID-19 infection in SSA, and the correlation coefficient between the weekly average number of confirmed COVID-19 cases reported by each country and the weekly average stringency index of its anti-COVID-19 policy measures.

**Results:** 84.61% (33) and 100% (39) of the 39 SSA countries for which predictions were made did not reach a total of 1 000 and 10 000 confirmed COVID-19 cases at the predicted interval dates. The daily number of confirmed COVID-19 cases was lower than the one projected for all SSA countries. The case fatality ratio of the COVID-19 infection in SSA was 3.42%. Among the 44 SSA countries for which the correlation could be estimated, it was negative for 17 (38.6 %) of them.

**Conclusions:** The natural characteristics of SSA and the public health measures implemented might partly explain that the actual data were lower than the predictions on the COVID-19 pandemic in SSA, but the low case ascertainment and the numerous asymptomatic cases did significantly influence this observation.

## INTRODUCTION

The Coronavirus disease 2019 (COVID-19) outbreak was classified as a pandemic on March 11^th^, 2020, owing to its lightning proliferation and the ominous prospect that the sudden massive influx of critically ill patients would overwhelm healthcare systems (1). Thenceforward, the world has experienced several COVID-19 surges and as of the April 30^th^, 2022, the planet has cumulatively reported 513 109 654 confirmed cases and 6 260 020 deaths (2). The pandemic has caused substantial disruptions in daily activities and has wreaked havoc socioeconomic systems across the globe.

The first case of COVID-19 infection on the African continent was detected in Egypt on February 12^th^, 2020 (3,4). As of April 30^th^, 2022, African countries have recorded a cumulative total of 11 895 452 confirmed COVID-19 cases and 253 791 deaths (2). At the onset of the COVID-19 pandemic in sub-Saharan Africa, the rapid progression of the sanitary crisis constrained local stakeholders to constantly make several important policy decisions on a short notice. Historical data could not be used to this effect given the novelty of the COVID-19 infection. Therefore, to raise alarms and build public health strategies, public health officials and stakeholders had to rely on predictive models. The latter had predicted that the COVID-19 pandemic would severely affect Sub-Saharan Africa (SSA), owing to its high levels of air traffic with China, the predominance of ill-equipped and short-staffed hospitals, the lack of research and testing laboratories, and the presence of unhealthy populations who live mostly in overcrowded urban centers with limited access to handwashing, but who interact through physical contacts-based sociocultural living customs (5). In the light of these concerning prospects, the World Health Organization (WHO) alerted on April 17^th^, 2020 that Africa would become the next epicenter of the COVID-19 pandemic (6,7).

Nevertheless, after the first waves of the COVID-19 pandemic had subsided, several reports highlighted that SSA had defied these dire predictions (8). However, to date, no study has verified this observation. More importantly, the world is currently experiencing a resurgence of COVID-19 cases driven by new and more virulent viral strains (9,10). It is therefore critical to scientifically evaluate if SSA did truly defy the grim forecasts on the COVID-19 pandemic, because if that were the case, it would be urgent to identify the population and environmental characteristics as well as the public health interventions which contributed to that positive outcome, as they would form the core of efficient strategies which would abate current and future COVID-19 occurrences. The proposed research undertook that endeavor; we compared the predictions against the actual data on the spread and burden of the COVID-19 pandemic in sub-Saharan Africa, and we discussed the potential reasons for the differences observed.

## METHODS

### Data sources

We conducted a cross-sectional study which compared predictions to actual data on the spread and burden of COVID-19 in SSA during the period spanning from March 1^st^ to September 30^th^, 2020. The predictions on the burden of COVID-19 in SSA were obtained from the MRC Centre for Global Infectious Disease Analysis at Imperial College London (11). The MRC Centre for Global Infectious Disease Analysis at Imperial College London used data from the European Centre for Disease Control database to predict for each country within the next 28 days: [a] the daily number of confirmed COVID-19 cases; [b] the number of deaths; [c] the number of individuals needing oxygen or mechanical ventilation; and [d] the impact of changing the current intervention policy (11). The source of the predictions on the spread of COVID-19 in SSA was a publication by Pearson et al. from the Center for Mathematical Modeling of Infectious Diseases of the London School of Hygiene & Tropical Medicine (1). Pearson et al. predicted the 95% confidence interval (CI) dates at which each SSA country would report a total of 1 000 and 10 000 COVID-19 cases (1). The starting point of their predictions was the day when each country had at least 25 COVID-19 cases reported in the World Health Organization (WHO) Situation Reports (SITREPs); for countries who had not reached this number by the March 22^nd^, 2020, they used the number of cases reported in the WHO SITREPs as of March 23^rd^, 2020, 10:00 Central European Summer Time (1).

The WHO SITREPs are reports based on actual COVID-19 data provided by the health authorities of each country (12). Each SITREP presents the following: [a] the daily number of new confirmed COVID-19 cases; [b] the total number of confirmed COVID-19 cases reported since the date that a country started sending reports; [c] the number of days since the last day that a country reported a COVID-19 case; [d] the daily number of deaths attributed to COVID-19; [e] the total number of deaths attributed to COVID-19 since the date that the country started sending reports; and finally, [f] the updated COVID-19 transmission classification level of each country (level 1 = no case reported, level 2 = sporadic cases, level 3 = clusters of cases, and level 4 = community transmission) (12). From the WHO SITREPs, we extracted the actual dates at which each SSA country reported 1 000 and 10 000 confirmed COVID-19 cases. In instances where these exact numbers were unavailable, we retrieved the earliest dates at which they were surpassed. In instances where these numbers were never reached, we presented the total number of confirmed COVID-19 cases reported at the date this paper was submitted for peer review and publication. Of note, Pearson et al. did not compute predictions for Botswana, Burundi, Comoros, Guinea-Bissau, Malawi, Mali, Lesotho, Sao-Tome & Principe, and Sierra Leone (1), whereas they all belong to SSA. In a sake of completeness, we retrieved the total number of confirmed COVID-19 cases reported by these countries on April 30^th^ and May 31^st^, 2020, respectively. These dates were chosen because Person et al. had predicted that most SSA countries would report a total of 1 000 COVID-19 cases by the end of April and 10 000 cases couple of weeks later (1). In line with our primary objective of comparing predictions to actual data, we further scavenged the WHO SITREPs to find the actual dates at which these specific countries reported these numbers of cases.

To complement our research, we also evaluated the impact of the anti-COVID-19 policy measures implemented. The latter were valued through the Stringency Index (SI). The SI is a metric conceived as part of the Oxford COVID-19 Government Response Tracker (OxCGRT) project and has been previously described in details (13). In brief, the OxCGRT project is an endeavor undertaken by Oxford University to measure the strictness of the anti-COVID-19 policy measures that were enacted by the authorities of each country. To provide a general appreciation of each country’s governance, the OxCGRT project performed multiple different combinations of several individual policy measures to create a myriad of distinct indices. Among these indices, the SI combines nine of the response measures: school closures, workplace closures, cancellation of public events, restrictions on public gatherings, closures of public transport, stay-at-home requirements, public information campaigns, restrictions on internal movements, and international travel controls. Based on its level of enforcement, each of the response measures was given a score, and these individual scores were added together to yield the SI of each country. The SI was proportional to the strictness of each country’s anti-COVID-19 response, and a SI of 100 indicated the strictest response.

### Data Analyses

To compare predictions to actual data on the spread of COVID-19, we assessed if the actual dates at which each SSA country reached a total of 1 000 and 10 000 COVID-19 cases belonged to the respective 95% CI dates predicted by Pearson et al. (1). A country was said to have defied the predictions if the actual date did not belong to the predicted 95% CI. To compare predictions and actual data on the burden of COVID-19, we first performed a graphical comparison between the predicted and confirmed daily number of COVID-19 cases, from the day that each country first reported COVID-19 cases in the WHO SITREPs until September 30^th^, 2020.

Then, we calculated the case fatality ratio of the COVID-19 infection (proportion of deaths among all the identified confirmed cases of COVID-19 (14)) in SSA during the period spanning from March 1^st^ to September 30^th^, 2020. To evaluate the impact of the anti-COVID-19 policy measures implemented, we computed the correlation coefficient between the weekly average SI of anti-COVID-19 policy measures at time t, and the weekly average number of confirmed COVID-19 cases at time t + 14 days, along with the 95% CI of the said correlation coefficient. We considered a 14-day lag between the 2 metrics because it takes approximately one incubation period to see the effects of newly implemented anti-COVID-19 policy measures (15). Further, we used weekly averages instead of daily numbers to account for the delays in case ascertainment and reporting. Finally, to prevent any violation of the assumption of independence required for any correlation analyses, we used the repeated measures correlation technique available via the rmcorr package of the statistical software R (16). Of note, our report aligns with the Strengthening The Reporting of Observational studies in Epidemiology (STROBE) guidelines (Annex 1) (17).

### Ethics

Our study was exempt from Institutional Review Board approval since we used data already collected and published.

## RESULTS

### Predictions versus actual data on the spread of COVID-19

Among the 39 countries included in the publication by Pearson et al. (1) 33 (84.61%) did not reach a total of 1 000 COVID-19 cases at the predicted interval dates (table 1a). Of these, three (7.70%) countries reached 1000 confirmed cases only in 2021 (Mauritius: 31 March 2021, Seychelles: 23 January 2021, and Tanzania: 2 August 2021). All (100%) of the 39 countries included in the publication by Pearson et al. [1] did not reach a total of 10 000 COVID-19 cases at the predicted interval dates (table 1a). No (0%) country reported 10 000 cases before the predicted interval dates, 16 (41%) countries reached 10 000 cases only in 2021, and as of April 30^th^, 2022, 4 (10.25%) countries still have not reached 10 000 confirmed COVID-19 cases (Chad: 7411 cases, Eritrea: 9734 cases, Niger: 7434 cases, and Liberia: 8928 cases). Of the 9 countries not included in the publication by Pearson et al. (1), none (0%) of them either reported a total of 1000 on April 30^th^, 2020 or 10 000 COVID-19 cases on May 31^st^, 2020(table 1b). As of April 30^th^, 2022, 4 (44.44%) of these countries still have not reached 10 000 confirmed COVID-19 cases (Comoros: 8100 cases, Guinea-Bissau: 8202 cases, Sao Tome & Principe: 5957 cases, and Sierra Leone: 7681 cases).

**Table 1:** Predictions versus actual data on the spread of COVID-19 in SSA. 1a. Predicted and actual dates at which sub-Saharan African countries included in the publication of Pearson et al.,^1^ reported a cumulative total of 1000 and 10 000 confirmed COVID-19, respectively.

| Country | Actual date for 1000 cases<br>(number of cases) <sup>a</sup> | Predicted interval date for 1000<br>cases <sup>b</sup> | Actual date for 10 000 cases<br>(number of cases) <sup>c</sup> | Predicted interval date for 10 000<br>cases <sup>d</sup> |
| --- | --- | --- | --- | --- |
| Angola | 29 Jul 2020 (1000) | 12 Apr 2020 – 17 May 2020 | 28 Oct 2020 (10 074) | 28 Apr 2020 – 12 Jun 2020 |
| Benin | 26 Jun 2020 (1017) | 08 Apr 2020 – 16 May 2020 | 18 Aug 2021 (10 183) | 24 Apr 2020 – 09 Jun 2020 |
| Burkina Faso | 06 Jul 2020 (1000) | 31 Mar 2020 – 15 Apr 2020 | 24 Jan 2021 (10 038) | 14 Apr 2020 – 01 May 2020 |
| Cabo Verde | 26 Jun 2020 (1003) | 11 Apr 2020 – 16 May 2020 | 18 Nov 2020 (10,000) | 27 Apr 2020 – 11 Jun 2020 |
| Cameroon | 18 Apr 2020 (1017) | 27 Mar 2020 – 17 Apr 2020 | 19 Jun 2020 (10 638) | 12 Apr 2020 – 03 May 2020 |
| CAR | 01 Jun 2020 (1011) | 07 Apr 2020 – 14 May 2020 | 14 Jun 2021 (10 047) | 23 Apr 2020 – 09 Jun 2020 |
| Chad | 27 Aug 2020 (1004) | 11 Apr 2020 – 18 May 2020 | 30 April 2022 (7411) | 28 Apr 2020 – 12 Jun 2020 |
| Congo | 23 Jun 2020 (1087) | 06 Apr 2020 – 13 May 2020 | 06 Apr 2021 (10 084) | 22 Apr 2020 – 06 Jun 2020 |
| Côte d’Ivoire | 24 Apr 2020 (1004)* | 02 Apr 2020 – 02 May 2020 | 03 Jul 2020 (10 244) | 16 Apr 2020 – 24 May 2020 |
| DRC | 11 May 2020 (1024) | 01 Apr 2020 – 19 Apr 2020 | 29 Aug 2020 (10 007) | 15 Apr 2020 – 06 May 2020 |
| Djibouti | 26 Apr 2020 (1008)* | 11 Apr 2020 – 18 May 2020 | 13 Apr 2021 (10 077) | 27 Apr 2020 – 12 Jun 2020 |
| Equatorial Guinea | 24 May 2020 (1043) | 06 Apr 2020 – 10 May 2020 | 9 Sept 2021 (10 284) | 21 Apr 2020 – 04 Jun 2020 |
| Eritrea | 27 Dec 2020 (1039) | 11 Apr 2020 – 17 May 2020 | 30 April 2022 (9734) | 02 May 2020 – 11 Jun 2020 |
| Eswatini | 07 Jul 2020 (1011) | 07 Apr 2020 – 15 May 2020 | 04 Jan 2021 (10 138) | 23 Apr 2020 – 10 Jun 2020 |
| Ethiopia | 31 May 2020 (1063) | 04 Apr 2020 – 01 May 2020 | 20 Jul 2020 (10 207) | 18 Apr 2020 – 26 May 2020 |
| Gabon | 14 May 2020 (1004) | 06 Apr 2020 – 10 May 2020 | 18 Jan 2021 (10 019) | 21 Apr 2020 – 05 Jun 2020 |
| Gambia | 08 Aug 2020 (1090) | 10 Apr 2020 – 19 May 2020 | 12 Dec 2021 (10 034) | 28 Apr 2020 – 12 Jun 2020 |
| Ghana | 20 Apr 2020 (1042) | 01 Apr 2020 – 17 Apr 2020 | 11 Jun 2020 (10 201) | 15 Apr 2020 – 03 May 2020 |
| Guinea | 27 Apr 2020 (1094)* | 06 Apr 2020 – 15 May 2020 | 12 Sep 2020 (10 020) | 22 Apr 2020 – 09 Jun 2020 |
| Kenya | 21 May 2020 (1029) | 03 Apr 2020 – 24 Apr 2020 | 13 Jul 2020 (10 105) | 18 Apr 2020 – 12 May 2020 |
| Liberia | 13 Jul 2020 (1010) | 08 Apr 2020 – 14 May 2020 | 30 April 2022 (7434) | 24 Apr 2020 – 09 Jun 2020 |
| Madagascar | 07 Jun 2020 (1026) | 08 Apr 2020 – 25 Apr 2020 | 29 Jul 2020 (10 104) | 21 Apr 2020 – 11 May 2020 |
| Mauritania | 09 Jun 2020 (1049) | 06 Apr 2020 – 14 May 2020 | 09 Dec 2020 (10 105) | 23 Apr 2020 – 07 Jun 2020 |
| Mauritius | 31 Mar 2021 (1028) | 06 Apr 2020 – 01 May 2020 | 30 August 2021 (10 196) | 19 Apr 2020 – 25 May 2020 |
| Mozambique | 07 Jul 2020 (1012) | 13 Apr 2020 – 19 May 2020 | 11 Oct 2020 (10 001) | 29 Apr 2020 – 14 Jun 2020 |
| Namibia | 17 Jul 2020 (1032) | 05 Apr 2020 – 13 May 2020 | 17 Sep 2020 (10 078) | 22 Apr 2020 – 07 Jun 2020 |
| Niger | 17 Jun 2020 (1016) | 11 Apr 2020 – 19 May 2020 | 30 April 2022 (8928) | 28 Apr 2020 – 12 Jun 2020 |
| Nigeria | 25 Apr 2020 (1095) | 23 Mar 2020 – 18 Apr 2020 | 01 Jun 2020 (10 162) | 09 Apr 2020 – 04 May 2020 |
| Rwanda | 30 Jun 2020 (1001) | 02 Apr 2020 – 22 Apr 2020 | 13 Jan 2021 (10 122) | 16 Apr 2020 – 11 May 2020 |
| Senegal | 02 May 2020 (1024) | 23 Mar 2020 – 12 Apr 2020 | 31 Jul 2020 (10 106) | 07 Apr 2020 – 29 Apr 2020 |
| Seychelles | 23 Jan 2021 (1033) | 05 Apr 2020 – 08 May 2020 | 25 May 2021 (10 740) | 20 Apr 2020 – 02 Jun 2020 |
| Somalia | 11 May 2020 (1054)* | 06 Apr 2020 – 12 May 2020 | 21 Mar 2021 (10 085) | 27 Apr 2020 – 06 Jun 2020 |
| South Africa | 28 Mar 2020 (1170)* | 26 Mar 2020 – 15 Apr 2020 | 11 May 2020 (10 015) | 09 Apr 2020 – 03 May 2020 |
| South Sudan | 09 May 2020 (1111)* | 05 Apr 2020 – 14 May 2020 | 25 Mar 2021 (10 048) | 22 Apr 2020 – 08 Jun 2020 |
| Togo | 07 Aug 2020 (1001) | 30 Mar 2020 – 19 Apr 2020 | 30 Mar 2021 (10 249) | 15 Apr 2020 – 05 May 2020 |
| Uganda | 09 Jul 2020 (1000) | 10 Apr 2020 – 08 May 2020 | 14 Oct 2020 (10 069) | 24 Apr 2020 – 31 May 2020 |
| Tanzania | 02 Aug 2021 (1017) | 06 Apr 2020 – 02 May 2020 | 10 October 2021 (25 846)<sup>#</sup> | 20 Apr 2020 – 26 May 2020 |
| Zambia | 28 May 2020 (1057) | 09 Apr 2020 – 15 May 2020 | 19 Aug 2020 (10 218) | 25 Apr 2020 – 11 Jun 2020 |
| Zimbabwe | 14 Jul 2020 (1034) | 11 Apr 2020 – 17 May 2020 | 01 Dec 2020 (10 129) | 28 Apr 2020 – 11 Jun 2020 |

**Table 1:** Predictions versus actual data on the spread of COVID-19 in SSA. 1b. Total number of confirmed COVID-19 cases reported on 30 April 2020 and 31 May 2020 by the sub-Saharan African countries which were not included in the publication of Pearson et al.,^1^ and dates at which these countries reported a cumulative of total of 1000 and 10 000 confirmed COVID-19 cases in the World Health Organization Situation Reports.

| Country | Actual cases as of 30 April 2020 <sup>a</sup> | Date for 1000 cases <sup>b</sup> | Actual cases as of 31 May 2020 <sup>c</sup> | Date for 10 000 cases <sup>d</sup> |
| --- | --- | --- | --- | --- |
| Botswana | 23 | 08 Oct 2020 (1066) | 35 | 26 Nov 2020 (10 258) |
| Burundi | 11 | 11 Jan 2021 (1019) | 63 | 16 August 2021 (10 103) |
| Comoros | 01 | 07 Jan 2021 (1050) | 106 | 30 April 2022 ( 8100) |
| Guinea-Bissau | 205 | 18 May 2020 (1032) | 1256 | 30 April 2022 ( 8202) |
| Lesotho | 0 | 21 Aug 2020 (1015) | 02 | 15 Feb 2021 (10 254) |
| Malawi | 37 | 26 Jun 2020 (1005) | 284 | 15 Jan 2021 (11 223) |
| Mali | 490 | 23 May 2020 (1015) | 1265 | 31 Mar 2021 (10 042) |
| ST & P | 14 | 07 Dec 2020 (1002) | 483 | 30 April 2022 ( 5957) |
| Sierra Leone | 124 | 08 Jun 2020 (1001) | 861 | 30 April 2022 ( 7681) |

<sup>a</sup> Originates from the World Health Organization (WHO) situation reports (SITREPs) and is the cumulative total of confirmed COVID-19 cases reported by each country as of 30 April 2020. <sup>b</sup> Originates from the WHO SITREPs and it is the date at which each country reported a cumulative total of 1000 confirmed COVID-19 cases ; in instances where this exact number could not be found is reported the earliest date at which this number was surpassed, and in instances where this number was never reached is reported the date at which this paper was submitted for peer review and publication. <sup>c</sup> Originates from the WHO SITREPs and is the cumulative total of confirmed COVID-19 cases reported by each country as of 31 May 2020. <sup>d</sup> Originates from the WHO SITREPs and it is the date at which each country reported a cumulative total of 10 000 confirmed COVID-19 cases ; in instances where this exact number could not be found is reported the earliest date at which this number was surpassed, and in instances where this number was never reached is reported the date at which this paper was submitted for peer review and publication. ST & P = Sao Tome & Principe.

**Table 2.** Impact of the anti-COVID-19 policy measures implemented. Repeated measures correlation between the weekly average number of confirmed COVID-19 cases reported and the weekly average stringency index of the anti-COVID-19 policy measures, along with the 95% confidence interval of the correlation coefficient, using data spanning from the first day that each county reported cases in the World Health Organization Situation Reports until September 30th, 2020.

| Country | Correlation coefficient <sup>a</sup> | 95% confidence interval of the correlation coefficient |  | P-value |
| --- | --- | --- | --- | --- |
|  |  | Upper bound | Lower bound |  |
| Angola | - 0.025 <sup>b</sup> | - 0.46 | 0.42 | 0.91 |
| Benin | 0.27 | - 0.19 | 0.63 | 0.21 |
| Botswana | - 0.04 <sup>b</sup> | - 0.48 | 0.42 | 0.87 |
| Burkina Faso | - 0.32 <sup>b</sup> | - 0.65 | 0.13 | 0.14 |
| Burundi | 0.23 | - 0.25 | 0.62 | 0.31 |
| Cabo Verde | 0.20 | - 0.26 | 0.60 | 0.36 |
| Cameroon | -0.03 <sup>b</sup> | - 0.45 | 0.40 | 0.90 |
| Central African Republic | 0.21 | - 0.24 | 0.60 | 0.33 |
| Chad | 0.75 | 0.48 | 0.90 | <0.0001 |
| Comoros |  | ** <sup>c</sup> |  |  |
| Congo | 0.004 | - 0.43 | 0.43 | 0.90 |
| Côte d'Ivoire | -0.04 <sup>b</sup> | - 0.46 | 0.40 | 0.86 |
| Democratic Republic of Congo | 0.30 | - 0.15 | 0.64 | 0.17 |
| Djibouti | 0.27 | - 0.17 | 0.63 | 0.20 |
| Equatorial Guinea |  | ** <sup>c</sup> |  |  |
| Eritrea | - 0.083 <sup>b</sup> | - 0.51 | 0.40 | 0.71 |
| Eswatini | 0.22 | - 0.23 | 0.60 | 0.30 |
| Ethiopia | 0.002 | - 0.43 | 0.43 | 0.99 |
| Gabon | 0.09 | - 0.36 | 0.50 | 0.70 |
| Gambia | - 0.18 <sup>b</sup> | - 0.57 | 0.27 | 0.40 |
| Ghana | - 0.04 <sup>b</sup> | - 0.50 | 0.40 | 0.84 |
| Guinea | 0.22 | - 0.23 | 0.60 | 0.31 |
| Guinea-Bissau |  | ** <sup>c</sup> |  |  |
| Kenya | 0.07 | - 0.37 | 0.49 | 0.74 |
| Lesotho | 0.10 | - 0.44 | 0.60 | 0.72 |
| Liberia | 0.28 | - 0.17 | 0.63 | 0.20 |
| Madagascar | 0.002 | - 0.44 | 0.44 | 0.90 |
| Malawi | 0.08 | - 0.40 | 0.51 | 0.72 |
| Mali | 0.34 | - 0.12 | 0.70 | 0.12 |
| Mauritania | 0.20 | - 0.26 | 0.60 | 0.40 |
| Mauritius | - 0.17 <sup>b</sup> | - 0.60 | 0.28 | 0.42 |
| Mozambique | 0.03 | - 0.41 | 0.46 | 0.90 |
| Namibia | 0.40 | - 0.08 | 0.70 | 0.08 |
| Niger | 0.60 | 0.20 | 0.81 | 0.004 |
| Nigeria | 0.23 | - 0.21 | 0.60 | 0.30 |
| Rwanda | 0.01 | - 0.42 | 0.44 | 0.90 |
| Sao-Tome & Principe |  | ** <sup>c</sup> |  |  |
| Senegal | 0.02 | - 0.40 | 0.44 | 0.91 |
| Seychelles | - 0.43<sup>b</sup> | - 0.73 | -0.007 | 0.04 |
| Sierra Leone | - 0.06 <sup>b</sup> | - 0.50 | 0.40 | 0.80 |
| Somalia | 0.33 | - 0.11 | 0.70 | 0.11 |
| South Africa | - 0.06 <sup>b</sup> | - 0.47 | 0.40 | 0.80 |
| South Sudan | - 0.11 <sup>b</sup> | - 0.55 | 0.40 | 0.70 |
| Tanzania | - 0.20 <sup>b</sup> | - 0.57 | 0.27 | 0.40 |
| Togo | - 0.12 <sup>b</sup> | - 0.52 | 0.32 | 0.60 |
| Uganda | -0.20 <sup>b</sup> | - 0.60 | 0.30 | 0.40 |
| Zambia | 0.41 | - 0.03 | 0.71 | 0.06 |
| Zimbabwe | -0.03 <sup>b</sup> | - 0.44 | 0.44 | 0.9 |

<sup>a</sup> Repeated measures correlation coefficient between the daily number of confirmed COVID-19 cases reported by each country and the stringency index of its anti-COVID-19 policy measures. <sup>b</sup> Repeated measures correlation coefficient indicating a negative correlation between the weekly average number of confirmed COVID-19 cases reported by each country and the weekly average stringency index of its anti-COVID-19 policy measures. <sup>c</sup> The repeated measures correlation coefficient between the weekly average number of confirmed COVID-19 cases and the weekly average stringency index anti-COVID-19 policy measures as well as its 95% confidence interval were not estimated because data on the stringency index for these countries are not available in the Oxford Coronavirus Government Response Tracker (OxCGRT)

### Predictions versus actual data on the burden of COVID-19

From March to September 2020, except for Burundi, Botswana, and Seychelles, the actual daily number of confirmed COVID-19 cases reported by each SSA country was lower than the number predicted (figure 1). At the continental level, the total number of actual confirmed COVID-19 cases was 1 126 341, which is far lower than the total number of predicted cases (median 10 504 027 IQR: 9 936 984 – 11 170 105). The cumulative case fatality ratio of the COVID-19 infection in SSA from March to September 2020 was approximately 3.42%.

**Figure 1:**
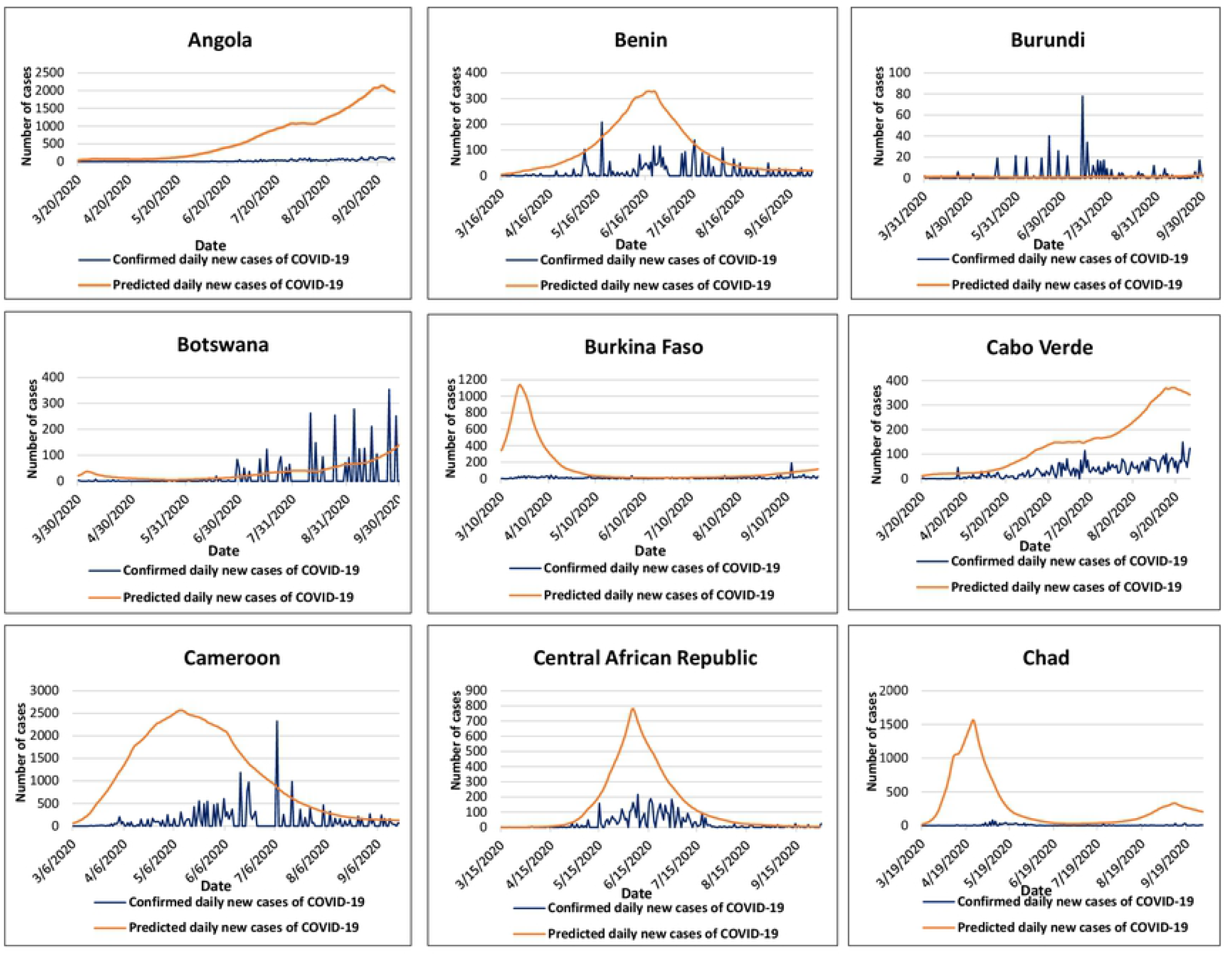
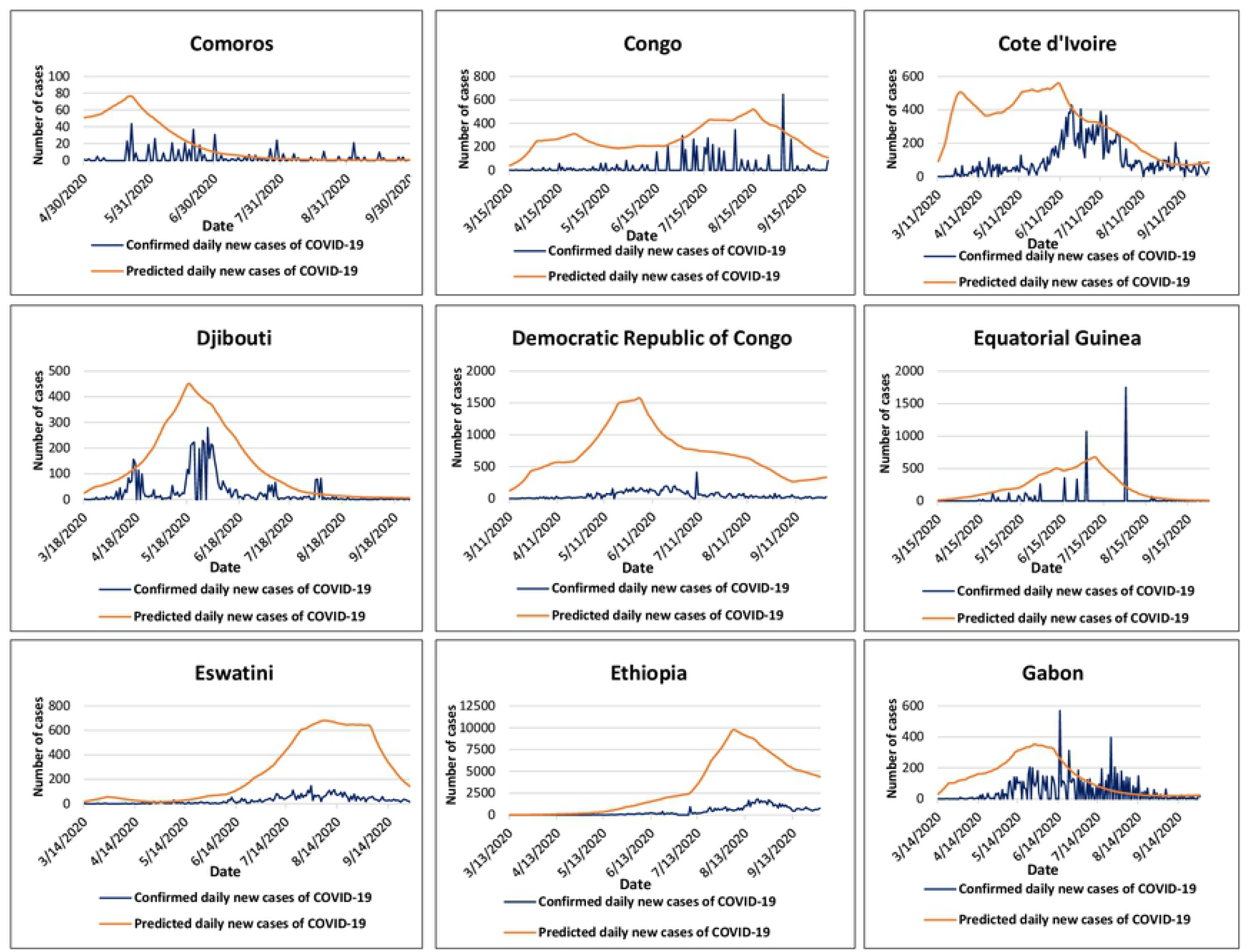
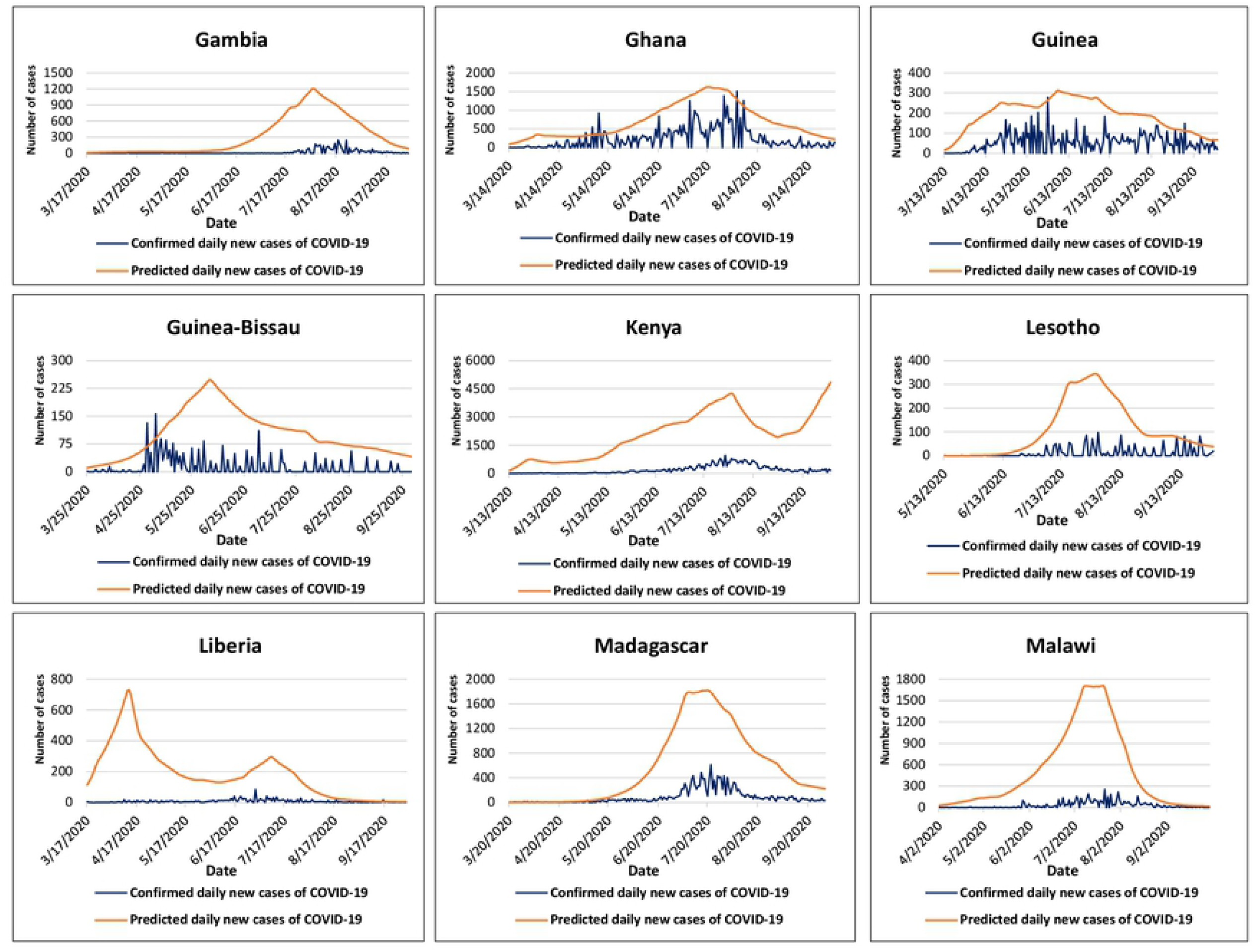
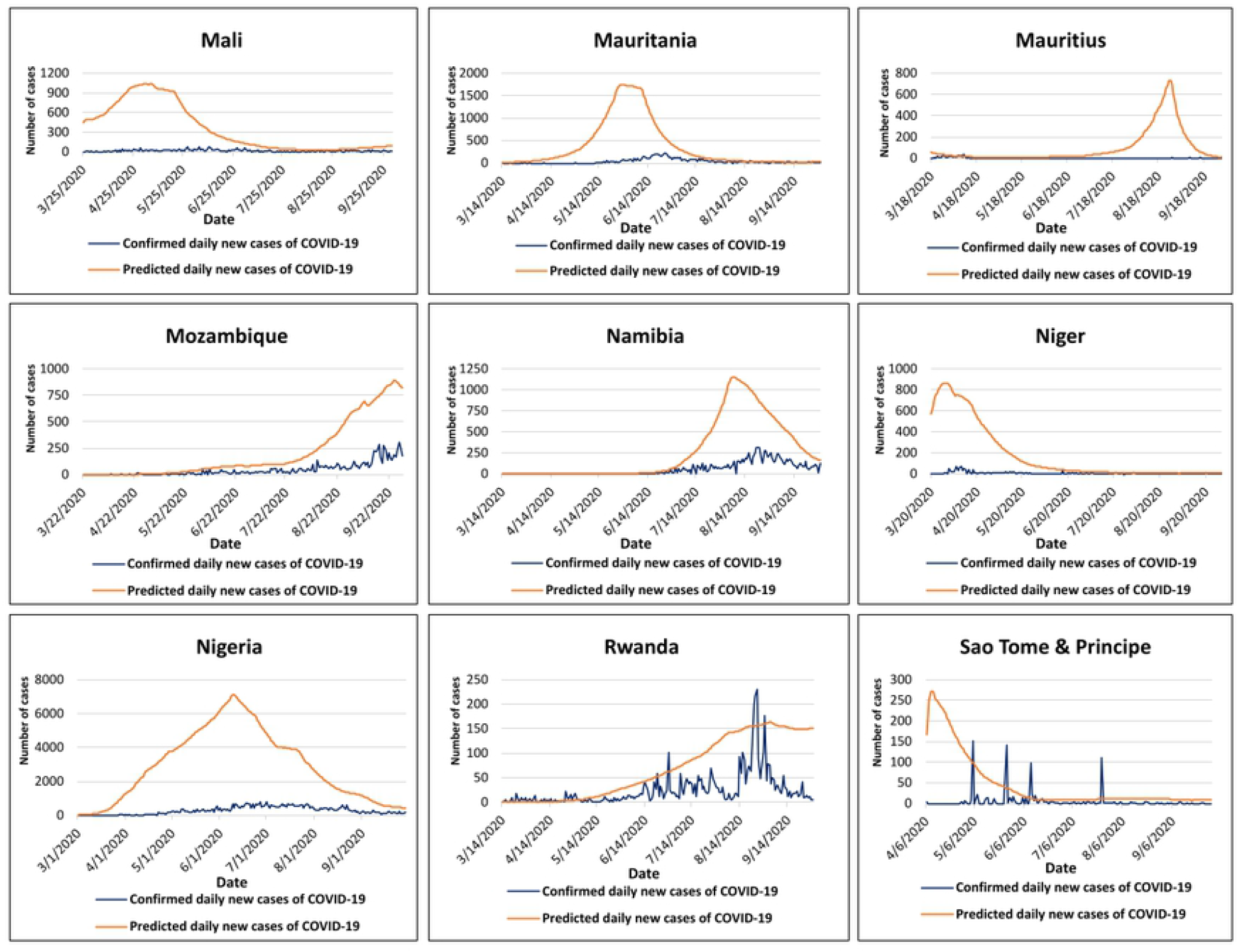
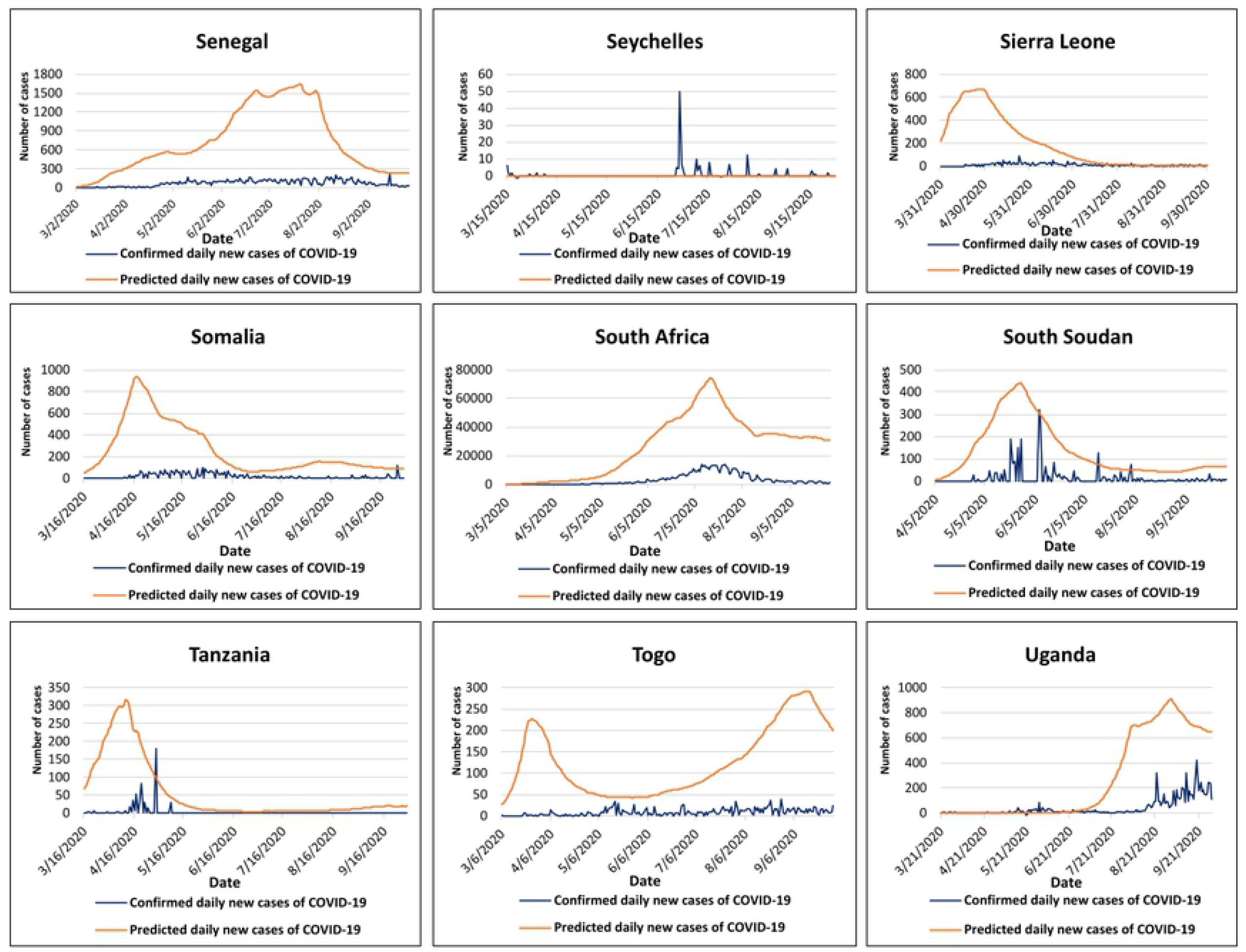
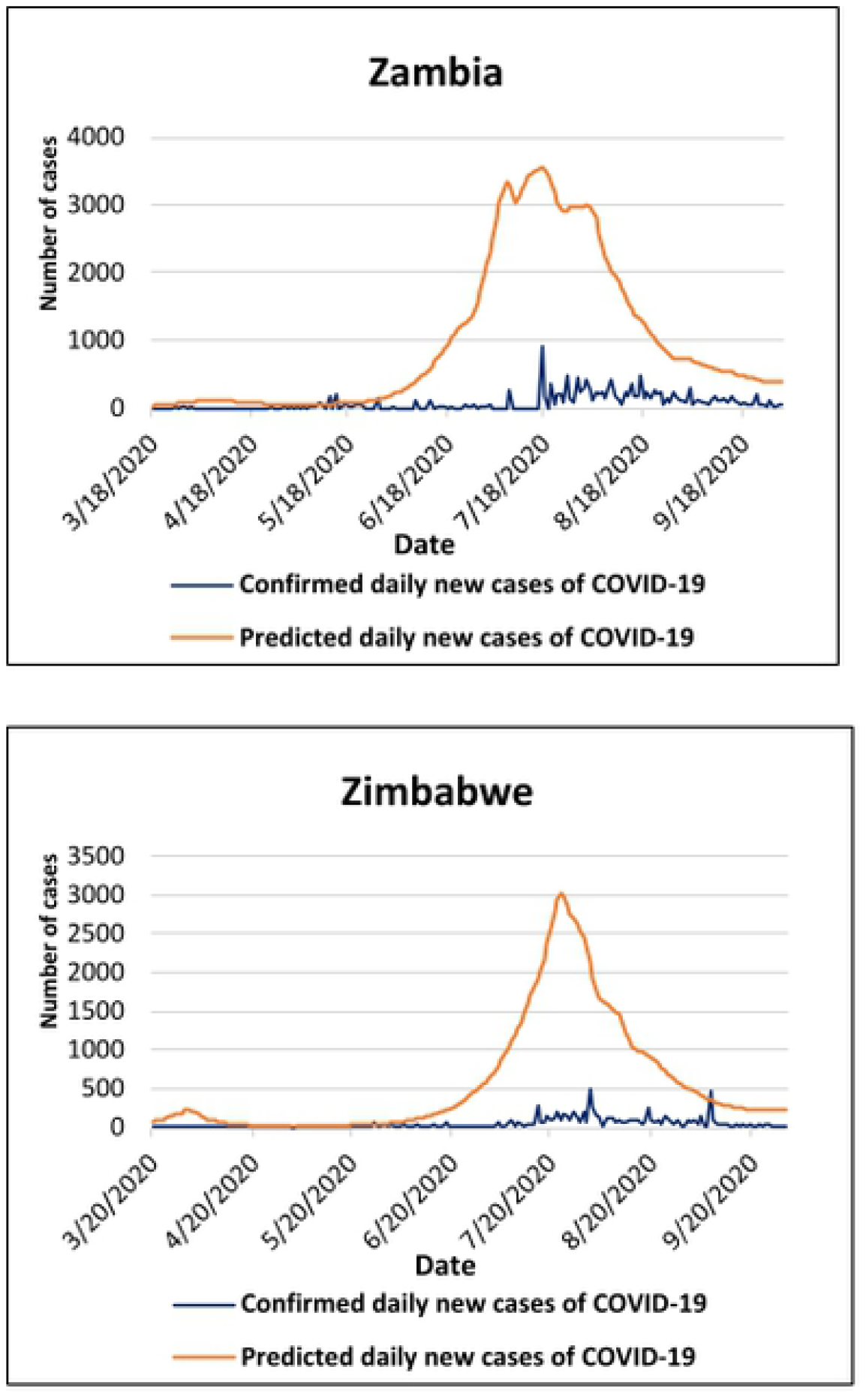
The burden of the first wave of the Coronavims Disease 19 (COVID-19) in sub Saharan Africa and other continents. **la**. Comparison between predicted and confirmed daily new cases of COVID-19 in sub-Saharan African countries, from the day that each country first reported COVID-19 cases in the World Health Organization Situation Reports, to September 30^th^, 2020. **Source:** World Health Organization Situation Reports ^6^; The MRC Centre for Global Infectious Disease Analysis at lmperial College London. ^*5*^

### Impact of the anti-COVID-19 policy measures implemented

Among the 44 countries for which the correlation could be calculated, a negative correlation was found for 17 (38.6%) of them, and none of these negative correlation coefficients had a 95% CI which excluded 0. The correlation coefficient for Comoros, Equatorial Guinea, Guinea-Bissau, and Sao Tome & Principe were not estimated because data on the stringency index for these countries are not available in the Oxford Coronavirus Government Response Tracker.

## DISCUSSION

Overall, most SSA countries did not report 1 000 and 10 000 cases at the predicted dates, and the actual numbers of COVID-19 cases were lower than those predicted. These results might be explained by the limitations of the statistical models which yielded these predictions. Additionally, specific local population and environmental characteristics as well as the low case ascertainment might have had a mitigating effect.

The prediction model of the MRC Centre for Global Infectious Disease Analysis at Imperial College London was built on estimates of severity obtained from data from China and Europe, and model parameters obtained from data from China and the United Kingdom (11). On the other hand, Pearson et al. considered that the reproductive number R (which is the number of ancillary cases that one case would generate if in contact with a completely susceptible population (18)) would be 2, that the dispersion estimate k (which is the variance of R over the mean of R and quantifies whether a set of observed cases are clustered or dispersed when compared to cases following a standard negative binomial distribution (19)) would be 0.58, and that the serial interval (which is the time that elapses between two consecutive cases of an infectious disease (20)) would be normally distributed with a mean of 4.7 ± 2.9 days (1). These model parameters all originated from populations which substantially differ from SSA populations in terms of composition, density, living customs, and health status, all of which impact the dynamic of the COVID-19 pandemic. Furthermore, Abbot et al. acknowledged that data were scarce at the time they estimated R and k, and they pointed that their results would be significantly impacted if new data became available (21). Lastly, Rai et al. also indicated that their study calculating the serial interval had “numerous limitations”, including the high risk of bias due to the multiplicity of data collection protocols, the impossibility to identify every potential contact an individual had, and the numerous unaccounted asymptomatic travelers (22).

Pearson et al. also assumed that no public health interventions would be implemented (1). However, as of March 31^st^, 2020, more than 50% of SSA countries had already imposed travel restrictions to prevent the importation of the SARS-CoV-2 virus (23). Of note, previous evidence had reported that travel bans were effective in preventing the importation of COVID-19 (24,25). Pearson et al. also assumed that both the early epidemic trends and the reporting fraction among actual cases and delay would remained constant, that there would always be sufficient unreported infections to continue the transmission, and that new cases would represent a sample from both identified and previously unidentified transmission chains (1). Nevertheless, for such assumptions to be true, the living conditions and the dynamic of the COVID-19 infection must have also remained unchanged. However, living conditions were and are still constantly modified to match the momentum of the COVID-19, and the pandemic is an ever-changing outbreak which outsmarts all assumptions and predictive models.

In SSA, specific population and environmental characteristics also mitigated the epidemic pace of the COVID-19 pandemic which was determined by the rate of introduction of the SARS-CoV-2 virus, its environmental proliferation, and the maintenance of the COVID-19 infection (14).

Evidence have demonstrated that the SARS-CoV-2 virus was introduced in SSA via international flights and tourists’ arrivals (14). Data show that 71% (34 out 48) of SSA countries had imposed travel bans on flights arriving from high-risk areas for COVID-19 several days before (indicated in green and as negative numbers in figure 2a) the first confirmed COVID-19 case(s) was(were) identified. As a result, tourists’ arrivals significantly decreased from March to June 2020 and culminated at a record −91% in April 2020 compared to the same month in 2019 (figure 2b). Previous evidence had demonstrated that non-pharmacologic interventions of this type can abate the spread of the COVID-19 infection (26). Indeed, efficient bureaucracy and guidance happening prior to high infection rates is known to have produced the greatest benefits against COVID-19 (14). Therefore, these early interventions potentially mitigated the rate of introduction of the SARS-CoV-2 virus in SSA.

**Figure 2:**
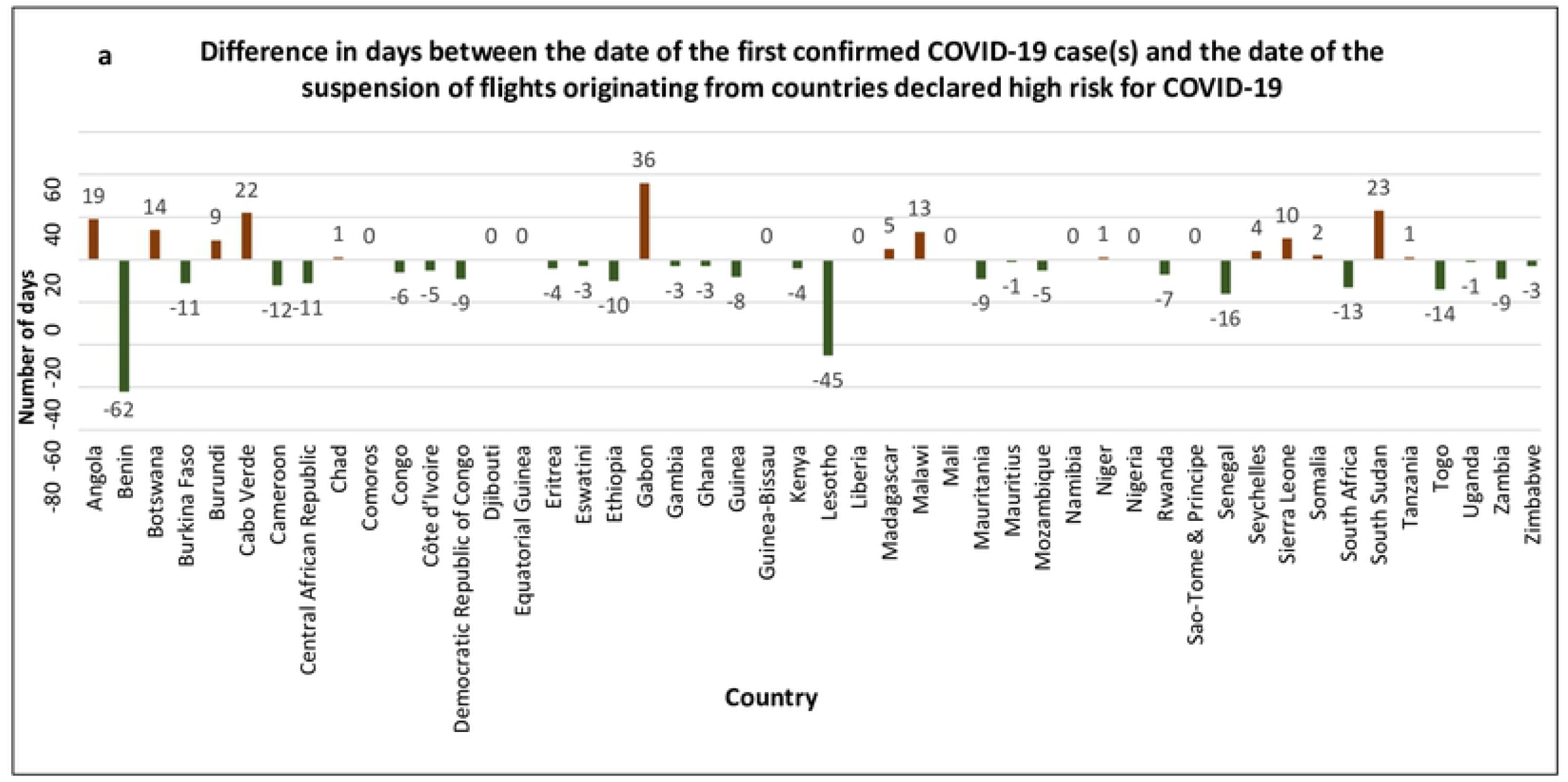
Public Health response of sub-Saharan African countries against COVID-19. **2a**: Number of days between the date of the first confirmed COVID 19 case(s) and the date at which the governments of sub-Saharan Afrcancounrries suspended flights originating from the countries which had been declared high risk for COVID 19. Data source: The Oxford COVID-19 Government Response Tracker(OxCGRT) project ^7^

**Figure 2:**
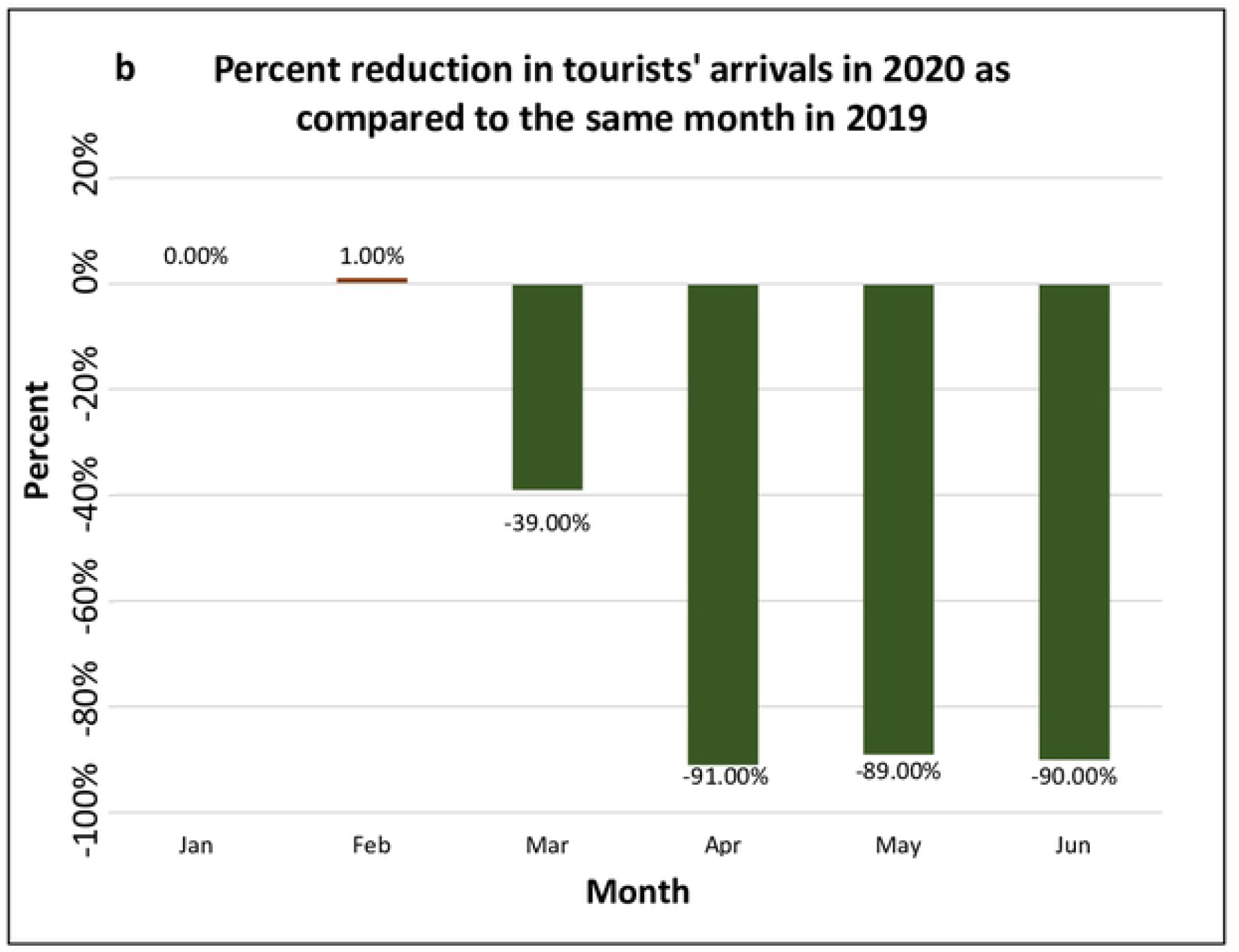
Public Health response of sub Saharan African countries against COVID 19. **2b**: Reduction in tourists’ arrivals in sub-Saharan African countries in 2020, expressed as the percentage difference in the number of tourists’ arrival in the same compared to the same month in 2019. Data source : The United Nations World Tourism Organization ^27^

The rate of introduction of the SARS-CoV-2 virus was also determined by the susceptibility of the exposed population. The Centers for Disease Control and Prevention (CDC) described that the people at high risk for COVID-19 infection were those of older age (above 65 years), and those with health risk factors such as obesity and/or severe medical conditions like diabetes (27). Population pyramids (figure 3a) built using data from the 2020 United Nations Population Prospects (28) show that subjects aged 0 to 14 years represent 40.55% of the SSA population, whereas those aged 65 and older represent only 3.5%. This is quite low compared to North America, Europe, and Asia where the 65 and older represent 16%, 18.8 % and 19.6% of the total population respectively. In addition, data compiled from the Global Burden of Disease Study 2017 (29) reveals that except for malnutrition, HIV/AIDS, and tuberculosis, SSA reported lower mortality rates from obesity, smoking, cardiovascular disease, chronic kidney disease, chronic respiratory diseases, chronic liver disease, cancers, diabetes, and outdoor pollution, compared to other continents where the COVID-19 pandemic was more severe (figure 3b). At last, it is hypothesized that SSA populations possess a cross-protection against COVID-19 because of anterior infections by epidemic coronaviruses or other germs (14,30,31).

**Figure 3:**
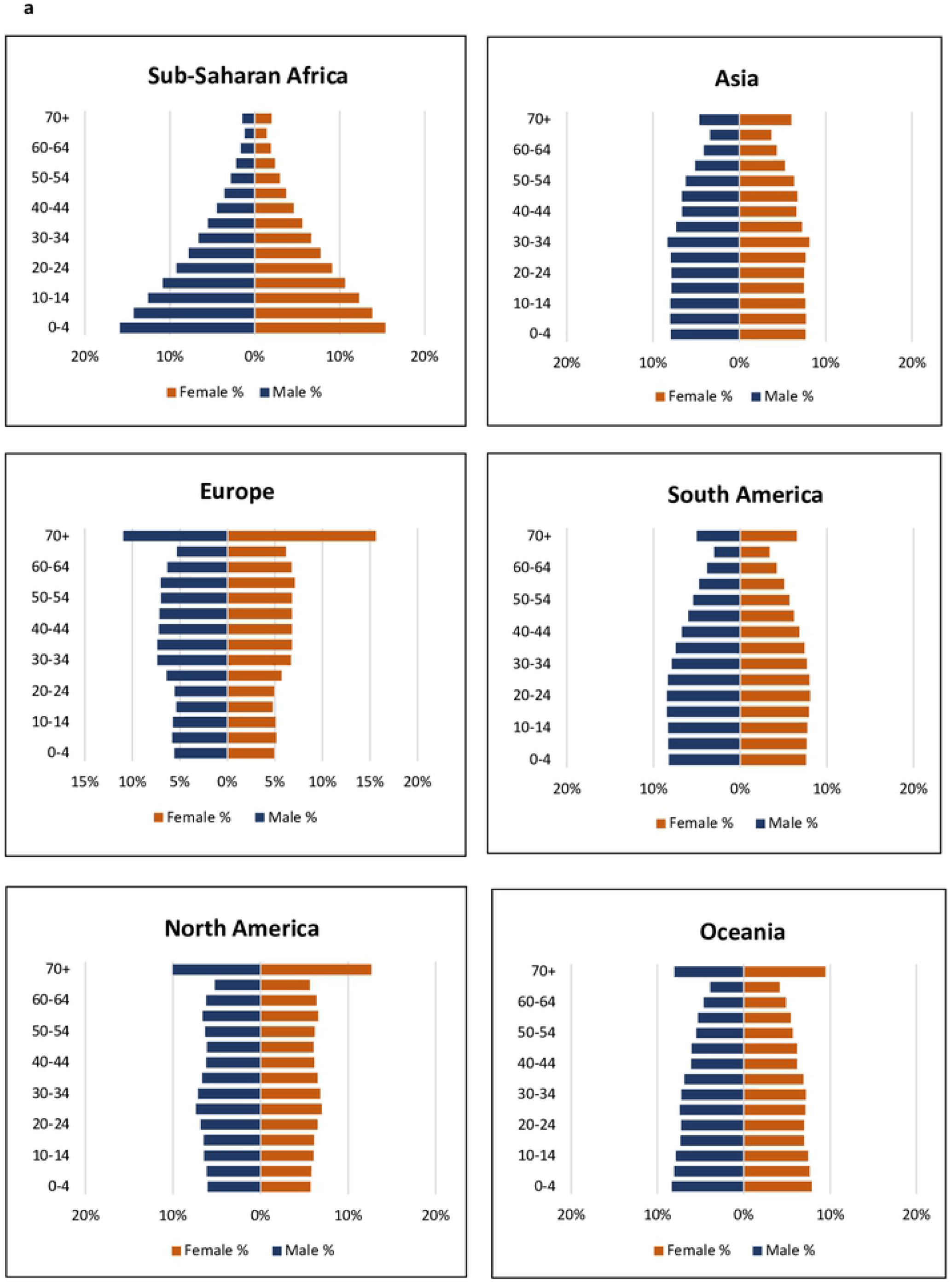
Susceptibility of populations to COVID-19. **3a**: Population pyrainids of sub-Saharan Africa, Asia, Europe, South America, North America, and Oceania. Source: The Global Burden of Diseases, Injuries, and Risk Factors Study (GBD) 2017 ^20^

**Figure 3:**
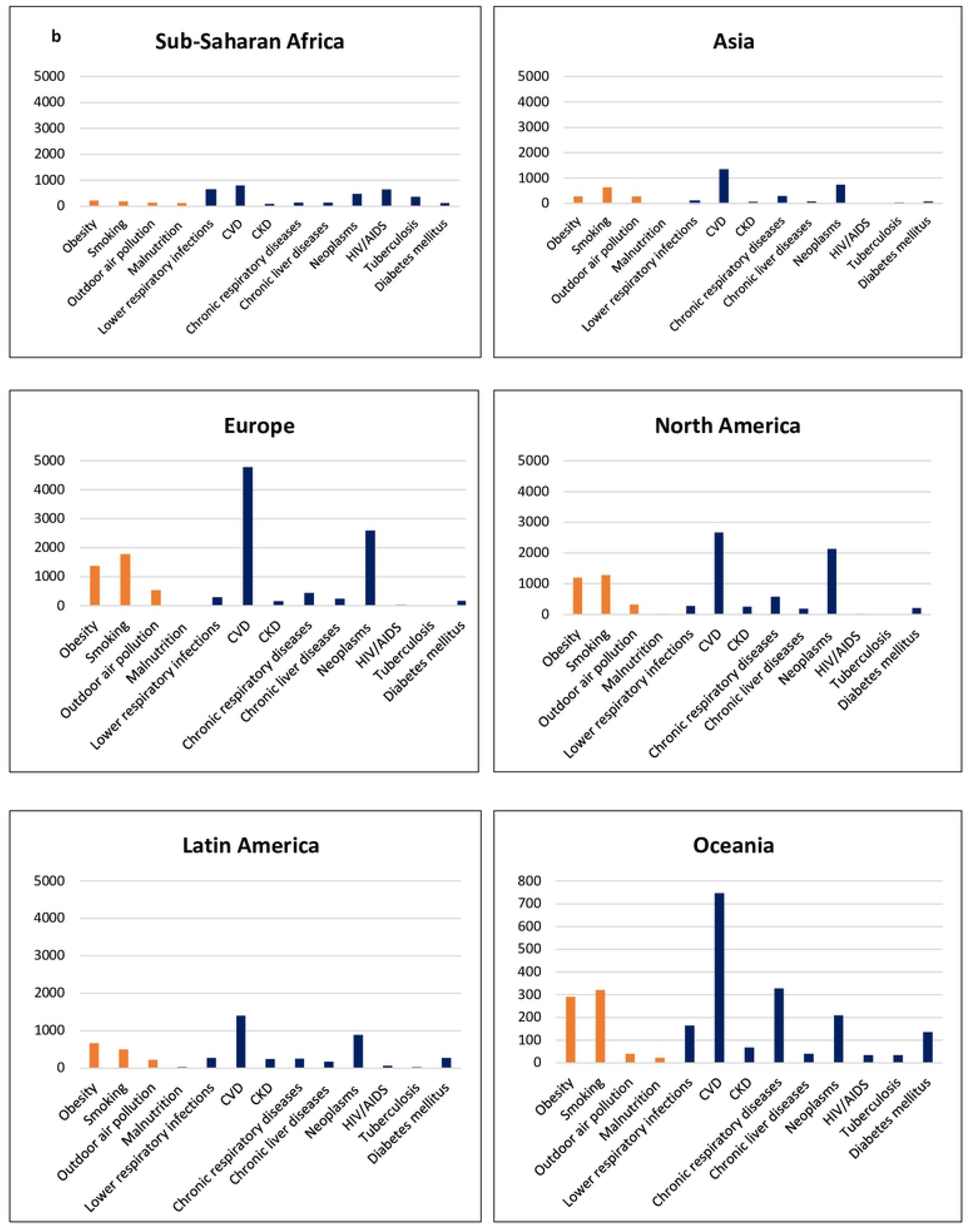
Susceptibility of populations to COVID 19. **3b**: Annual mortality per 100,000 inhabitants, from specific risk factors and chronic diseases, on the 6 continents. Source: The Global Burden of Diseases, Injuries, and Risk Factors Study (GBD) 2017 ^20^

Climate was also considered a major determinant of the COVID-19 pandemic in SSA. Indeed, it was advanced that the warmer and wetter weather of SSA tempered the environmental proliferation of the SARS-CoV-2 virus in the region (32). However, these evidence were not welcomed by all (14). This skepticism probably arose because the temperatures required to kill the SARS-CoV-2 virus are far beyond what humans can tolerate (33). Nevertheless, we still believe that the climate had a mitigating effect on the COVID-19 pandemic in SSA, not by a direct effect on the virus, but rather by an indirect effect on the immunity through the influence that climate has on Vitamin D (Vit-D) levels. Indeed, data from the Solar Atlas (34) shows that SSA is the region which receives the highest amount of solar energy, and this translates into high levels of Vit-D in local populations (35). In turn, Vit-D is known to significantly improve human immune capacities (36).

Despite these potential protective characteristics, SSA still reported COVID-19 cases which means that the virus was introduced in the region but did not spread at the rates predicted. This hints that the transmissibility of the COVID-19 infection among people was also affected. Non-pharmacologic interventions are known to abate the spread of COVID-19 (26), and the governments of SSA countries implemented preventive measures early on during the first wave of the pandemic (figures 2a and 2b). Other determinants of the transmissibility of the COVID-19 infection are the connectivity, the travel time, and the habitual population movements between cities and regions (14). During the last 50 years, the increase in road constructions across SSA significantly improved the connectivity and reduced the travel time between localities (14). However, the daily commute is not custom in SSA. Therefore, the spread of the COVID-19 infection might have been limited to narrow geographic areas. The transmissibility of the COVID-19 infection is also impacted by the prevalence of severe cases. Indeed, severe cases are more prone to transmit the COVID-19 infection because of their higher viral load, and they are also more prone to die of the infection (37). The case fatality ratio of COVID-19 in SSA was approximately 3.42%, which is much lower compared than the case fatality ratio reported on other continents (38–40).

Evidence suggest that the lower susceptibility of SSA populations to COVID-19 might have resulted in most cases being asymptomatic (41), meaning less contagious. Therefore, SSA might have hosted mostly less severe cases, which translated into a lower probability of transmission of the COVID-19 infection in the region.

The actual number of COVID-19 cases in SSA might also be this low because not all cases were reported. Nations with highly technical and infrastructural abilities were first to possess diagnostic equipment for COVID-19, and they prioritized their local populations. Consequently, SSA countries, which do not possess such capabilities, were left on endless waiting lists for supply, which substantially limited their testing capacities. As evidence, statistics on the number of daily COVID-19 tests per 1 000 individuals show that on 30 June 2020 data were missing for several SSA countries (42). This is corroborated by serologic studies conducted in a limited number of SSA countries and which have reported a seroprevalence between 1.8% and 45.1% (43–46). The low frequency of COVID-19 case ascertainment in SSA might also be due to the high number of asymptomatic subjects. Indeed studies have suggested that the lower susceptibility to COVID-19 conferred by the youth of SSA populations resulted in most cases being asymptomatic (41), and hence they were not tested and thus not reported.

Our study presents certain limitations. The cross-sectional design did not allow us to capture the temporal relationship between the different indicators and the occurrence of COVID-19 cases, which precludes any discussion about causation. However, it would have been unethical to conduct a prospective study in which humans would have been purposefully deprived of life-saving preventive measures against COVID-19.

Therefore, a cross-sectional study was indicated. Another limitation is the small number of countries (17 out of 44) for which there was a negative correlation between the weekly average number of COVID-19 cases and the weekly average SI, which questions the true efficacy of the prevention efforts deployed inside SSA countries. Of note, the scientists who designed the index cautioned that it does not measure the actual level of implementation of the anti-COVID-19 policy measures which make the index (13). Nevertheless, studies had demonstrated the efficacy of non-pharmacologic measures against COVID-19, and the SI was the sole metric available in that regard for our use. Finally, the vast array of the potential contributors to the spread of COVID-19, the unconsidered peculiarities of each SSA country, and the use of continental level rather individual level health data, point to the necessity to exercise caution in the interpretation of these results which were pooled across several very dissimilar geographic and socioeconomic settings.

Our research does have several strengths. It responds to the long-lasting call for a comprehensive analysis of the differences between the predictions and the actual data on the COVID-19 pandemic in SSA. In addition, our analysis also included the countries for which predictions were not made. Finally, we discussed the population and environmental characteristics as well as the public health interventions which may have contributed to the positive outcome observed. Substantial efforts should be made to reinforce the preventive measures against COVID-19 in SSA. Besides, resources should be allocated to strategies aiming to maintain the baseline health of populations in the world in general, and in SSA in particular. Finally, the grim prospect of future similar public health crisis calls for a thorough upgrade of the quantity and quality of diagnostic, treatment, research, and drug development facilities across the globe.

## CONCLUSION

The actual figures of COVID-19 were lower than the predictions for all SSA countries, but the low case ascertainment and the numerous asymptomatic cases have greatly influenced this observation. Exploring the hypotheses suggested to understanding the reasons for more asymptomatic cases in SSA could help build stronger strategies to respond to future COVID-19 resurgences as well as other viral epidemics. Finally, there is an urgent need for a massive upscaling of diagnostic, treatment, and research capabilities in SSA and across the globe.

## Data Availability

Data will be made available by the corresponding author upon a signed data access agreement.

## Acknowledgments

We thank Professor Martin Tenniswood for his comments and suggestions.

## Competing interests

I have read the journal’s policy and the authors of this manuscript have the following competing interests: Simeon Pierre Choukem who is co-author of this paper is also an Academic Editor for PLOS ONE journal. This does not alter our adherence to PLOS ONE policies on sharing data and materials.

Further, the conclusions made herein are those of the authors and do not necessarily represent the position of the institutions to which they belong.

## Authors’ Contributions

CDFM and SPC conceptualized the idea. CDFM devised the study, conducted the analysis, interpreted the results, and wrote the first draft of the manuscript. YB, DNTA, TLTY, and SPC thoroughly reviewed the manuscript for data completeness, methodologic accuracy, and grammatical correctness. All authors had full access to the data in the study and accept responsibility to publish the results.

## Data Sharing

Data will be made available by the corresponding author upon a signed data access agreement.

---

**Annex 1.** STROBE Statement—Checklist of items that should be included in reports of ***cross-sectional studies***

|  | Item |  | Page |
| --- | --- | --- | --- |
|  | No | Recommendation | No |
| **Title and abstract** | 1 | (a) Indicate the study’s design with a commonly used term in the title or the abstract | 3 |
|  |  | (b) Provide in the abstract an informative and balanced summary of what was done and what was found | 2 |
| **Introduction** |  |  |  |
| Background/rationale | 2 | Explain the scientific background and rationale for the investigation being reported | 3 |
| Objectives | 3 | State specific objectives, including any prespecified hypotheses | 3 |
| **Methods** |  |  |  |
| Study design | 4 | Present key elements of study design early in the paper | 3 |
| Setting | 5 | Describe the setting, locations, and relevant dates, including periods of recruitment, exposure, follow-up, and data collection | 3-4 |
| Participants | 6 | (a) Give the eligibility criteria, and the sources and methods of selection of participants | 3 |
| Variables | 7 | Clearly define all outcomes, exposures, predictors, potential confounders, and effect modifiers. Give diagnostic criteria, if applicable | 3-4 |
| Data sources/<br>measurement | 8* | For each variable of interest, give sources of data and details of methods of assessment (measurement). Describe comparability of assessment methods if there is more than one group | 3-4 |
| Bias | 9 | Describe any efforts to address potential sources of bias | 3-4 |
| Study size | 10 | Explain how the study size was arrived at | 3-4 |
| Quantitative variables | 11 | Explain how quantitative variables were handled in the analyses. If applicable, describe which groupings were chosen and why | 3-4 |
| Statistical methods | 12 | (a) Describe all statistical methods, including those used to control for confounding | 3-4 |
|  |  | (b) Describe any methods used to examine subgroups and interactions | 3-4 |
|  |  | (c) Explain how missing data were addressed | NA |
|  |  | (d) If applicable, describe analytical methods taking account of sampling strategy | 3-4 |
|  |  | (e) Describe any sensitivity analyses | NA |
| Results |  |  |  |
| Participants | 13* | (a) Report numbers of individuals at each stage of study—eg numbers potentially eligible, examined for eligibility, confirmed eligible, included in the study, completing follow-up, and analysed | NA |
|  |  | (b) Give reasons for non-participation at each stage | NA |
|  |  | (c) Consider use of a flow diagram | NA |
| Descriptive data | 14* | (a) Give characteristics of study participants (eg demographic, clinical, social) and information on exposures and potential confounders | 3-4 |
|  |  | (b) Indicate number of participants with missing data for each variable of interest | NA |
| Outcome data | 15* | Report numbers of outcome events or summary measures | 3-4 |
| Main results | 16 | (a) Give unadjusted estimates and, if applicable, confounder-adjusted estimates and their precision (eg, 95% confidence interval). Make clear which confounders were adjusted for and why they were included | NA |
|  |  | (b) Report category boundaries when continuous variables were categorized | NA |
|  |  | (c) If relevant, consider translating estimates of relative risk into absolute risk for a meaningful time period | NA |
| Other analyses | 17 | Report other analyses done—eg analyses of subgroups and interactions, and sensitivity analyses | NA |
| **Discussion** |  |  |  |
| Key results | 18 | Summarise key results with reference to study objectives | 4 |
| Limitations | 19 | Discuss limitations of the study, taking into account sources of potential bias or imprecision. Discuss both direction and magnitude of any potential bias | 9 |
| Interpretation | 20 | Give a cautious overall interpretation of results considering objectives, limitations, multiplicity of analyses, results from similar studies, and other relevant evidence | 6-9 |
| Generalisability | 21 | Discuss the generalisability (external validity) of the study results | 9 |
| **Other information** |  |  |  |
| Funding | 22 | Give the source of funding and the role of the funders for the present study and, if applicable, for the original study on which the present article is based | 10 |

